# Exploring Pre-Exposure Prophylaxis (PrEP) modality preferences among Black cisgender women attending family planning clinics in Chicago

**DOI:** 10.1101/2023.11.16.23298643

**Authors:** Amy K. Johnson, Emily Ott, Eleanor E. Friedman, Amy Moore, Isa Alvarez, Agustina Pandiani, Catherine Desmarais, Sadia Haider

## Abstract

**Background:** Despite Pre-exposure prophylaxis’s (PrEP) demonstrated effectiveness, Black cisgender women continue to be at an elevated risk for HIV acquisition and uptake of daily oral PrEP is low in this population in the US. As advancements in PrEP delivery options continue, it is important to understand women’s acceptability of these additional options, specifically Black cisgender women, in order to inform uptake and adherence among this population at increased need of HIV prevention options.

**Setting:** A cross-sectional survey among Black cisgender women ages 13-45 (inclusive) attending women’s health clinics in Chicago, IL, prior to the approval of CAB-LA.

**Methods:** Descriptive statistics were used to describe the sample and bivariate analysis was used to detect differences between categorical and outcome variables using chi-square test. Responses to open-ended questions were thematically coded to explore Black cisgender women’s attitudes and preferences between the three methods of PrEP delivery including: vaginal ring, long-acting injectable, and a combined method that would prevent both pregnancy and HIV.

**Results:** In total, 211 cisgender women and adolescents responded to the survey. Both injections and combination pills were popular among participants, with 64.5% and 67.3% expressing interest in these forms of PrEP, respectively. The least popular method was the vaginal ring option, with 75.4% of respondents indicating that they would not consider using this modality. Overall, responses were not statistically different between the two surveys administered (Chi square p-values for injection PrEP method 0.66, combination PrEP method 0.93, and ring PrEP method 0.66) suggesting that the popularity of each method was not dependent on clinic location or age of participants.

**Conclusion:** This research provides important insights into the preferences and attitudes of different PrEP modalities among Black cisgender women. As different modalities continue to be approved for use among cisgender women, more research is needed to investigate the acceptability and preferences of these different modalities in order to improve uptake and adherence among this population.

## INTRODUCTION

Despite major advances in HIV prevention and treatment, racial and gender disparities in HIV/AIDS incidence continue to persist. Of the 36,801 new HIV cases in the U.S. in 2019, nearly 16% of all new HIV infections occurred among heterosexual women. In particular, Black cisgender women in the U.S. are disproportionately affected by HIV and although annual infections remained stable overall from 2015 to 2019 among this population, the rate of new HIV infections among Black women is 11 times that of white women and four times that of Latina women. Specific to Chicago, 85% of new HIV infections among heterosexual women in Chicago were among Non-Hispanic Blacks (Chicago Department of Public Health, 2016). This shows us that effective prevention methods are not adequately reaching people who could benefit most and underscores the need to develop and implement effective HIV prevention strategies for women, with a specific focus on advancing strategies among the Black community.

Approved by the Food and Drug Administration (FDA) in 2012 for adult populations and then in May 2018 for high-risk adolescents, pre-exposure prophylaxis (PrEP) is a promising biomedical prevention strategy that has the potential to reduce HIV infection among HIV negative populations who are at risk for acquisition (Hosek et al., 2016; Machado et al., 2017; Pilgrim et al., 2016). Despite previous studies demonstrating that oral HIV-PrEP can reduce HIV incidence among women who are adherent to PrEP, awareness and uptake is particularly low among Black women (Auerbach et al., 2015). Barriers to oral PrEP uptake and adherence include cost, the burden of taking a daily pill, and concerns about potential health effects (both long-term and short-term effects) and have led to an underutilization of PrEP among eligible groups (Golub et al., 2013; Krakower & Mayer, 2015; Wilton et al., 2015; Young & McDaid, 2014). Most importantly, studies have focused primarily on its use for MSM with a lack of data for cisgender women (Anderson et al., 2016).

Given the under-representation of cisgender women in PrEP research efforts (Bailey et al., 2017), there is an urgent need to better understand the unique factors that influence Black cisgender women’s uptake and acceptance of PrEP in order to curtail HIV-related health inequities in this population. In order to better understand women’s preferences for delivery methods of PrEP we conducted a survey of reproductive-aged women (ages 13-45 years) attending family planning clinics and explored HIV prevention behavior, awareness, and acceptability of PrEP. Using semi-structured interview format, we assessed attitudes and preferences across three different PrEP delivery strategies, including: vaginal ring, long-acting injectable, and a combined method that would prevent both pregnancy and HIV. Participants were also asked how these modalities compared to the daily pill option, since at the time of this study was conducted this modality was the only currently approved option. It is important to note, as of December 2021, the FDA approved the first injectable therapy for use in both adults and adolescents, cabotegravir long-acting injectable (CAB-LA) (LaPreze, 2022).

## METHODS

### Setting

To examine PrEP preferences among Black cisgender women in Chicago, we conducted a survey among patients at two care locations: University of Chicago Ryan Center and Planned Parenthood of Illinois family planning clinic. All participants completed informed consent procedures, and this study was approved by the Institutional Review Boards at the University of Chicago (IRB17-0984; IRB18-0901), and Lurie Children’s Hospital (IRB2017-1410). Data was collected between January to August of 2019.

### Study population

Eligibility was as follows: English-speaking, self-identify as African American and/or Black, 13-45 years old (inclusive), live in Chicago, and report recent sexual activity (within the last 6-12 months). All participants over 18 years completed oral informed consent prior to engaging in the study. Oral informed assent was obtained for participants under the age of 18 years and a parental waiver of consent for minor participants was granted to protect privacy of participants.

### Measures

Participants completed a quantitative survey followed by a brief semi-structured qualitative interview. All survey data were self-reported. The survey captured information about PrEP awareness, acceptability, barriers and facilitators to uptake, PrEP modality preferences and demographic and behavioral domains. We have previously described many of the questions used in this survey (Haider, et al. 2022; Johnson et al. 2020). Finally, participants were asked their opinions on other ways to take PrEP (i.e., long-acting injectable, vaginal ring, combined with a birth control pill) and how these different methods compared to the daily pill option, the only current FDA-approved method at the time of survey administration (e.g., “If a long-acting injectable (a shot that lasts a while, like the depo shot for birth control) version of PrEP was available, would you consider taking it? Why or why not? How does the injectable compare to a daily pill?”).

### Data Analysis

Descriptive statistics were used to describe the sample and bivariate analysis was used to detect differences between categorical and outcome variables using chi-square test. Quantitative analysis was conducted in SAS 9.4 (Cary, North Carolina). Responses to open-ended questions were audio recorded and transcribed verbatim; responses were thematically coded by the first author and themes were discussed with the study team to ensure consensus of code application.

## RESULTS

In total 211 cisgender women and adolescents responded to the survey. Responses were not statistically different between the survey location or age of participants (chi-square p-values for injection PrEP method 0.66, combination PrEP method 0.93, and ring PrEP method 0.66) suggesting that the popularity of each method was independent on location and age of participants.

Long acting injectables were a popular choice among participants with 64.5% expressing interest in this form of PrEP. Common themes that emerged from semi-structured responses in favor of injectable PrEP included not having to take a daily pill (86.8% of favorable responses), and general preference for an injection (11.0%). Most common themes for disinterest in an injectable form of PrEP included not liking shots (53.3% of negative responses) and having previously had a negative experience with Depo-Provera (21.3%). Similarly, combination pills that combined PrEP and birth control were also popular with 67.3% of participants willing to consider this method should it become available. Common thematic reasons why a combination pill was considered included a single pill with dual prevention effects (54.9% of favorable responses) general favourability (24.7%) and convenience (16.9%). Common reasons for disinterest included a desire to keep the prevention methods separate (34.6% of negative responses), general disinterest (23.2%) and currently trying to conceive or otherwise have no need for birth control (23.2%). Far less popular was the vaginal ring option, with 75.4% of respondents indicating that they would not consider using this modality if it became available. Favorable themes for vaginal ring PrEP delivery systems included not having to take a daily pill (67.3% of favorable responses) and general interest (13.2%). Common areas of disinterest included not wanting the ring in their body (60.4%) and general negative feelings about the ring (13.2%) (Table 1).

**TABLE 1.**
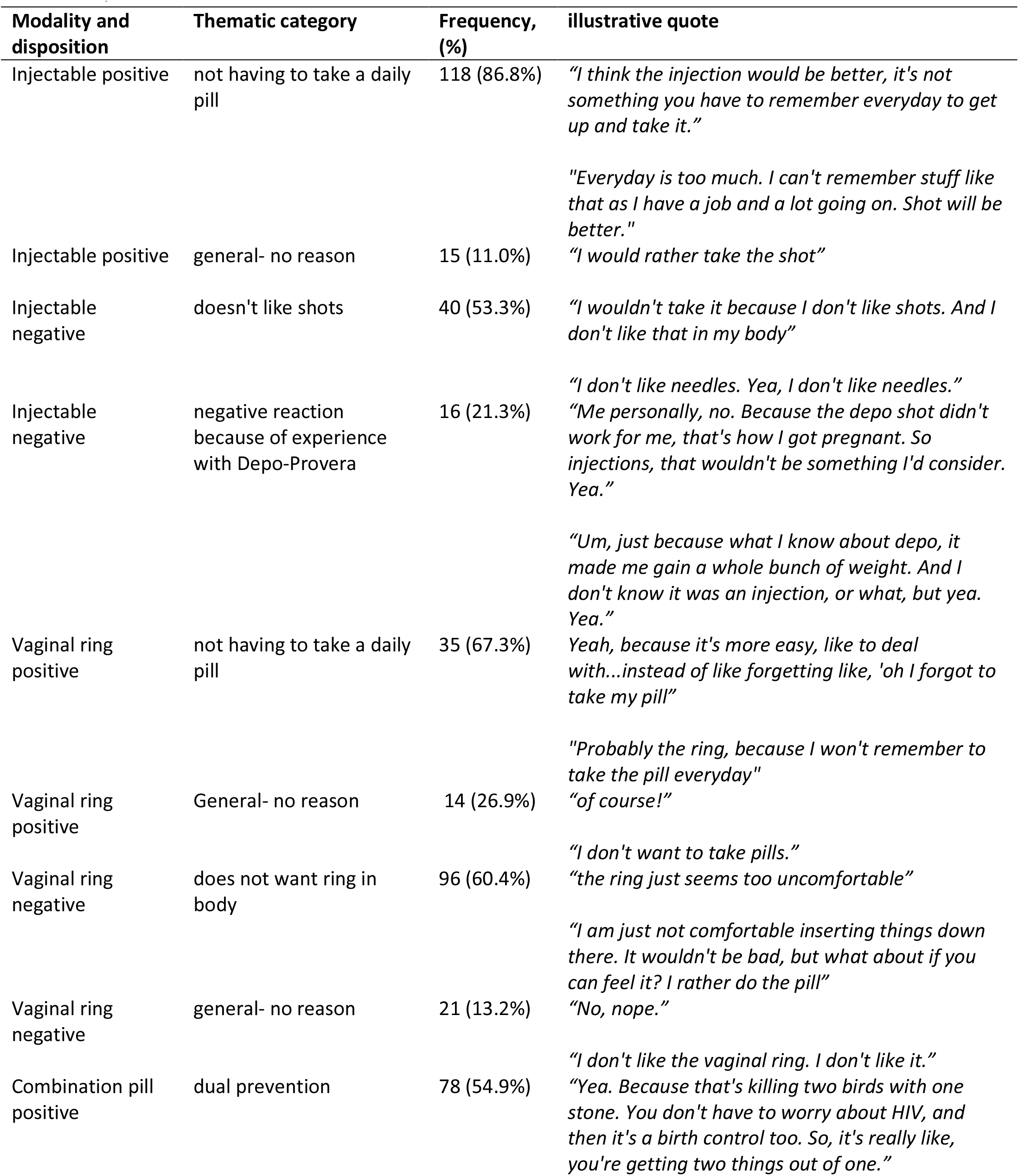
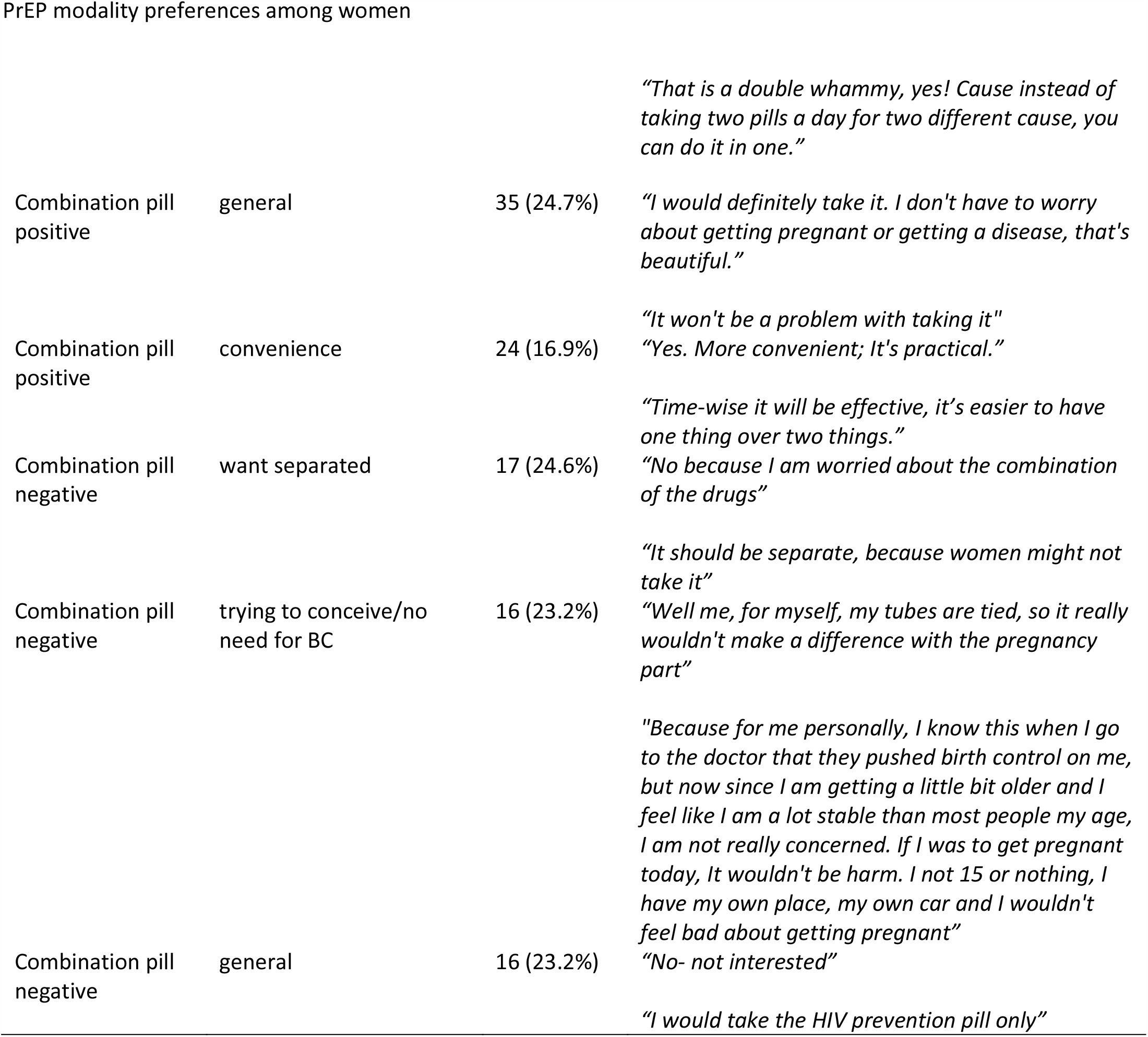
Qualitative Assessment of Themes Related to Modalities of PrEP.

Previous experiences, both positive and negative with birth control modalities also were distinctly listed as reasons for and against different PrEP modalities. Birth control pills, the Nuva ring, and Depo-Provera were all cited as explaining participants preferences for, or against similar delivery devices.

There was high overlap between those who were interested in combination and injection prevention methods (66.9%). For those interested in the ring method, high interest in both the injection and combination methods were seen (67.3% for both) but for those interested in either injection or combination methods, significantly lower interest in the ring method was seen (25.7%) (Table 2).

**TABLE 2.**
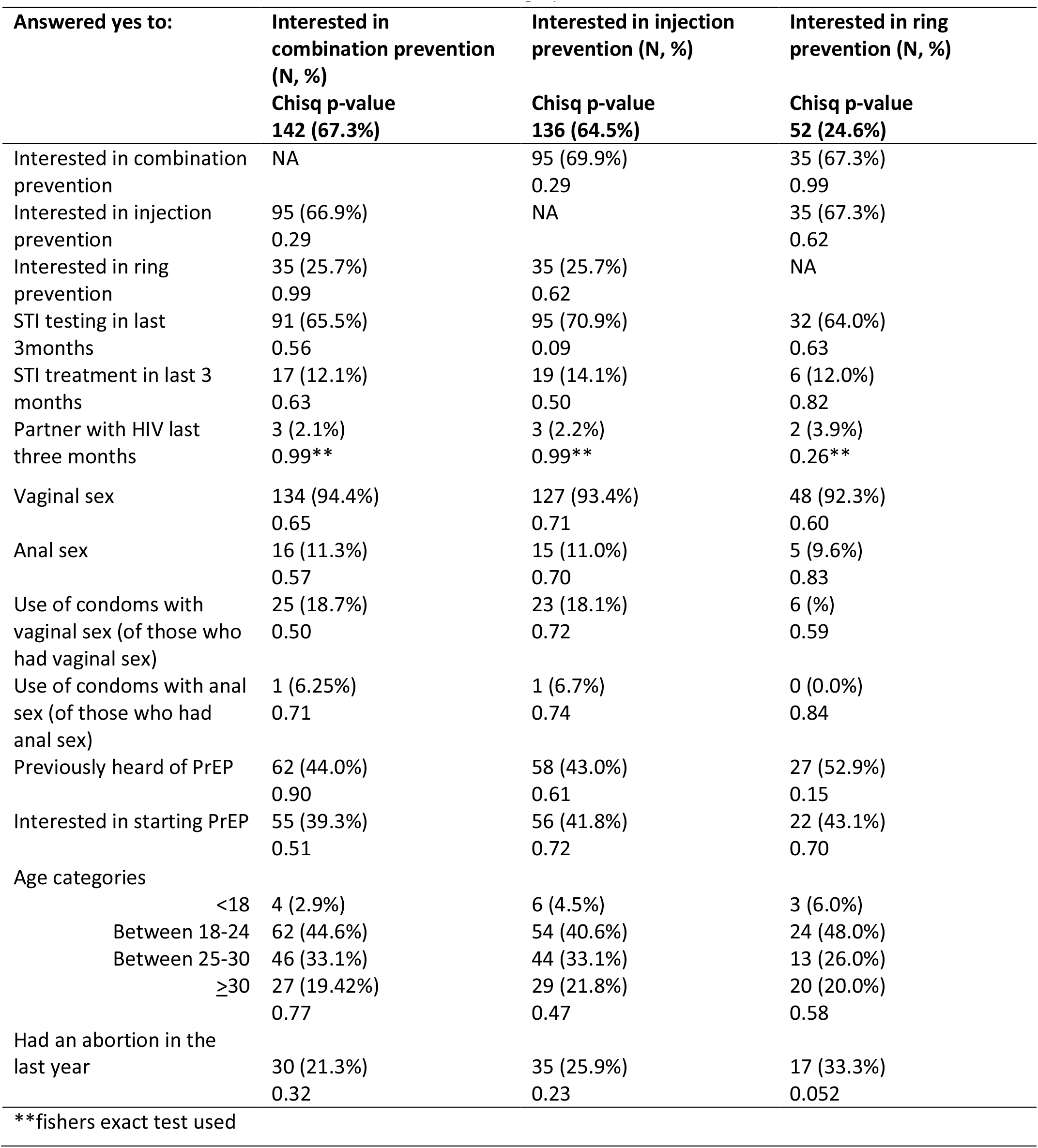
Modalities of PrEP and Associations with Demographic and Behavioral Characteristics.

When demographic and behavioral or sexual history factors were examined for associations with each PrEP modality no significant relationships were identified. Factors that were similar across preference for PrEP modalities included Sexually Transmitted Infection (STI) testing and treatment in the last three months, vaginal or anal sex in the last three months, use of condoms for either vaginal or anal sex, having heard of PrEP prior to the study and interest in starting PrEP (pill method). Participants who indicated that they were not interested in using PrEP as a daily pill did express interest in other forms of PrEP delivery, for example among those interested in the injection method, 37.0% of women originally expressed disinterest in using a separate PrEP pill. Similar results were seen for women interested in the combination pill but not the single use pill (40.3%), while fewer women were interested in the ring but not the daily separate pill (13.7%).

## DISCUSSION

HIV prevention efforts to date in the U.S., specifically PrEP scale-up initiatives, have not had a sufficient impact on uptake among Black cisgender women (Hodder et al., 2013). In the context of sustained rates of HIV among Black cisgender women and low uptake of daily oral PrEP, additional effective and desirable HIV prevention tools are needed. This study contributes data on Black cisgender women’s preferences for PrEP modalities, including the daily oral pill, LAI, ring, and combination methods. At the time this study was conducted the only approved method was the daily oral pill, so the other options, despite currently in development, were presented as theoretical options.

Popularity for each PrEP modality among Black cisgender women in our study were consistent with prior studies among women in the U.S. (Irie, 2022; Elopre, 2022). In our study, most women preferred injections (64.5%) and combination pills (67.3%) while majority of participants (75.4%) cited the vaginal ring as being their least preferred option. Based on responses in our study, women’s previous experiences with birth control methods were commonly cited as reasons for and against different PrEP modalities. Given the research on acceptability and implementation of contraceptive modalities over the past several decades (Delany-Moretlwe et al., 2016; Myers et al., 2015; Myers & Sepkowitz, 2013), it is possible that offering various delivery options of PrEP may improve uptake and adherence among ciswomen and perhaps even align with contraceptive preferences.

### Limitations

These results should be considered in light of the study’s limitations. First, our sample size is small which limited statistical power, variability in responses, and the inability to detect subgroup differences. Second, participants were recruited from two sexual health centers in an urban area and therefore our findings should not be interpreted as generalizable to Black cisgender women in totality. For instance, participants in this study may be better connected to sexual health information and as a result, have more knowledge and acceptability of PrEP overall compared to Black cisgender women not attending a sexual health center. Third, all data were self-reported and may be subject to social desirability; however, to mitigate socially desirable responses, quantitative data was collected via computer-assisted self-interviewing. Finally, at the time of survey administration participants were informed that the only currently approved and recommended form of PrEP was the daily oral pill. As stated above, in December 2021 (after the study was conducted), the FDA approved the first LAI PrEP (LaPreze, et al., 2022; FDA 2021). Had this information been available and known to participants at the time of survey administration, it is possible that acceptability and preferences toward the LAI PrEP modality may have been different. In addition, since we explored theoretical preferences for the various PrEP modalities and did not provide information about their relative costs or efficacy, our findings should be replicated in order to impact PrEP uptake.

## CONCLUSION

This study is one of the first of its kind to provide insight into preferences for and attitudes of PrEP modalities among Black cisgender women, including adolescent and young women. As advancements in PrEP delivery options continue, it is important to understand acceptability of additional options among subpopulations with disparate rates of HIV. Study findings have the potential to inform PrEP uptake and adherence as well as development, research, and clinical implementation for Black cisgender women.

## Data Availability

All data produced in the present study are available upon reasonable request to the authors

## Abbreviations

HIV: Human Immunodeficiency Virus
PrEP: Pre-exposure prophylaxis
STIs: Sexually Transmitted Infections
FDA: Food and Drug Administration
CAB-LAI: Cabotegravir long-acting injectable
MSM: Men who have sex with men

## Acknowledgements

The authors would like to thank the women who participated in the study and acknowledge Amber Olson, BS, for her assistance and support with recruitment.

